# Arterial Elasticity and Cardiac Function: A Cross-Sectional Study in the Vara- Skövde Cohort

**DOI:** 10.64898/2026.06.24.26356501

**Authors:** Gábor Szaló, Entela Bollano, Kristin Ottarsdottir, Karin Rådholm, Ying Li, Matthew Allison, Lyndia C Brumback, Margareta Hellgren, Ulf Lindblad, Bledar Daka

**Author notes:** Corresponding author: Gábor Szaló.

## Abstract

**Introduction:** Diastolic pulse wave analysis provides non-invasive estimation of arterial elasticity. Their clinical significance on the impairment of cardiac structure and function is not fully understood.

**Objective:** To examine associations between arterial elasticity assessed by diastolic pulse wave analysis and echocardiographic measures of cardiac structure and function in a community-based cohort.

**Methods:** A population-based cohort recruited 2816 randomly selected men and women aged 30–75 years between 2002 and 2005. A random subsample (n = 1,035) underwent echocardiography by a single senior cardiologist. Large-artery elasticity (C1) and small-artery elasticity (C2) were assessed by radial artery applanation tonometry. The analytical sample included 991 participants. Associations were examined using multivariable linear and logistic regression with sequential adjustment. The final model included sex, age, heart rate, diabetes mellitus, LDL cholesterol, body mass index, antihypertensive medication use, current smoking, alcohol intake, leisure-time physical activity, and systolic blood pressure.

**Results:** Among 991 participants, mean age was 51 years, 488 were men, and 160 had left ventricular hypertrophy. Mean C1 was 16.0 ± 5.1 mL/mmHg × 10, mean C2 was 6.9 ± 3.5 mL/mmHg × 100, and mean EF was 73.4 ± 8.5%. Higher C2 was associated with higher EF after systolic blood pressure adjustment (β per 1-SD increase: 1.2; 95% CI: 0.5–1.9; p < 0.001). Higher C1 was associated with lower odds of left ventricular hypertrophy (OR per 1-SD increase: 0.61; 95% CI: 0.44–0.84; p = 0.003).

**Conclusions:** Higher C2 was associated with better systolic function, whereas higher C1 was associated with lower odds of left ventricular hypertrophy.

**Clinical Perspective:** *What Is New?:* This study links non-invasive indices of arterial elasticity derived from diastolic pulse wave analysis to echocardiographic measures of cardiac structure and function in a community-based sample. Small-artery elasticity (C2) was positively associated with left ventricular ejection fraction, whereas large-artery elasticity (C1) was inversely associated with left ventricular hypertrophy, independent of cardiovascular risk factors and systolic blood pressure.

*What Are the Clinical Implications?:* Diastolic pulse wave analysis may provide clinically relevant insight into ventriculo–arterial interactions and cardiac remodelling. These findings suggest that early vascular assessment using diastolic pulse wave analysis may offer a non-invasive approach to identifying subclinical cardiovascular impairment before overt disease develops.

## Introduction

Heart failure (HF) is a major global health issue, contributing to high morbidity, mortality, and healthcare costs [1]. Echocardiography is central for the diagnosis and phenotyping of HF, providing detailed assessment of cardiac structure and function including left-ventricular systolic performance, diastolic function, and valvular disease [2]. Although HF is commonly classified based on ejection fraction (HFrEF: EF< 40%, HFmrEF: EF= 40–49%, HFpEF: EF ≥50%), the vascular and myocardial changes occur prior to clinical presentation.

Arterial stiffening, reflecting loss of arterial elasticity, is a key feature of vascular ageing and an important determinant of afterload and ventricular–arterial coupling [3]. Molecular mechanisms and vascular deterioration processes such as endothelial dysfunction, degradation of elastic fibres, increased collagen deposition, intimal and medial calcification are contributors to arterial stiffness [4]. Loss of arterial elasticity is a predictor of future hypertension, acute myocardial infarction, and stroke [5–9] and has also been investigated as a predictor of heart failure (HF) [10–12]. In HFpEF, arterial stiffness increases afterload and impairs ventricular-arterial coupling, leading to elevated filling pressures, higher pulse pressure, and concentric LV remodelling [3, 11].

Previous studies have shown that indices from diastolic pulse wave analysis predict incident HF and adverse cardiovascular outcomes [11–13]. To our knowledge, no previous study has examined diastolic pulse wave analysis and its potential association with cardiac function in the general population, despite its potential relevance to early pathophysiological changes. Subtle alterations in cardiac function may evolve gradually and may be associated with changes in cardiac structure and morphology linked to vascular dysfunction. Accordingly, the aim of this study was to investigate the association between vascular elasticity, assessed by diastolic pulse wave analysis, and cardiac function parameters measured by echocardiography.

## Methods

### Study population

The Vara–Skövde cohort was established to provide a broader perspective on the early detection of hypertension and impaired glucose metabolism, and to improve knowledge of potential preventive interventions. The baseline examination was conducted between 2002 and 2005. Participants were randomly selected from the population register, stratified by sex and five-year age groups, and included all individuals aged 30 to 74 years living in the municipalities of Vara and Skövde. No exclusion criteria were applied. A total of 2816 participants were enrolled at baseline, including 1400 men. Measurements of arterial elasticity were available for 2678 participants after exclusion of individuals with missing C1 or C2 data. Cardiac-function assessment was performed in a consecutively examined subsample of 1058 participants at baseline. Within this subsample, 67 participants were excluded because of incomplete data on arterial elasticity and/or cardiac function. The final cross-sectional analytical sample comprised 991 participants, including 488 men. No participants in the analytical study population had a previous diagnosis of heart failure or atrial fibrillation before the baseline examination.

#### Ethics approval and consent to participation

All participants were informed about the study procedures and objectives, and written informed consent was obtained from all participants. All methods were carried out in accordance with relevant guidelines and regulations. The study was approved by the Ethics Committee at the University of Gothenburg, Sweden (DNR: 199-01).

#### Measurements

Body size as height and weight were measured with participants wearing light clothing and no footwear, and values were rounded to the nearest 1 cm and 0.1 kg. Blood pressure was assessed in the right arm following a five-minute supine rest, and readings were recorded to the nearest 2 mmHg. Hypertension was diagnosed and classified according to the Joint National Committee 7 (JNC-7) guidelines [14]. All participants underwent a standard oral glucose tolerance test (OGTT) after a 12-hour overnight fast. Specifically, 75 g of glucose dissolved in water was ingested, and venous blood samples were obtained at baseline and 120 minutes post-load. Diabetes mellitus was diagnosed according to the 1999 World Health Organization criteria [15]. Laboratory analyses for LDL cholesterol and triglycerides were performed using standardized assays.

##### Large and Small Artery Elasticity Index

Arterial compliance was assessed using diastolic pulse contour analysis of the radial artery pressure waveform with the HDI/PulseWave™ CR-2000 system (Hypertension Diagnostics Inc., Eagan, MN, USA). After a standardized rest period (≥5 min), the right wrist was immobilized using a wrist stabilizer and applanations tonometer mounted in a holder was positioned over the site of maximal radial pulsation. Oscillometric brachial systolic and diastolic blood pressure were obtained using a cuff on the contralateral (left) arm and used to calibrate the radial waveform. Pressure waveforms were sampled at 200 Hz for 30 s per acquisition.

The device software fits a modified, third-order Windkessel model to the diastolic pressure decay and computes two elasticity indices: Large Artery Elasticity Index (C1/LAEI) reflecting large-artery (reservoir) compliance and Small Artery Elasticity Index (C2/SAEI) reflecting small-artery compliance, expressed as mL/mmHg. C1 and C2 are derived using device-estimated cardiac output, which is calculated from pulse waveform–derived timing variables and participant characteristics, including ejection time, heart rate, body surface area, and age.

##### Measurements of the cardiac function

All participants were examined by the same senior cardiologist, using standard two-dimensional transthoracic echocardiography (General Electrics VingMed S 5 System, operating with a 3.5 MHz-probe) [16, 17]. Echocardiographic images were acquired from parasternal and apical windows. Measurements for left ventricular (LV) calculations were taken from the Guidelines of the American Society of Echocardiography [18].

##### Assessment of Systolic function

Left ventricular systolic function was assessed in accordance with current guidelines. Ejection fraction (EF, %) was measured from apical four- and two-chamber views using a combination of the longitudinal systolic shortening mean index/atrioventricular plane displacement technique and a semi-quantitative visual estimation method [19, 20].

##### Left ventricular mass and left ventricular hypertrophy

LVM was calculated using the Devereux-modified ASE formula: LVM=0.8 × 1.04 [(LV internal dimension+posterior wall thickness+septal thickness)^3^− (LV internal dimension in diastole)^3^] + 0.6 g. LVM was then indexed for height^2.7^ (LVM-index, LVMI), and left ventricular hypertrophy (LVH) was defined as LVM-index ≥47.3 g/m^2.7^ based on guidelines [21, 22].

##### Lifestyles factors and medication

Information on medical history and lifestyle factors (such as alcohol consumption, smoking, and physical activity), were collected through validated questionnaires [23, 24]. Current smoking status was categorized as a binary variable (yes/no). Leisure-time physical activity levels were self-reported and provided four categorical options to describe the intensity and frequency of activity: sedentary, light physical activity, moderate physical activity, and vigorous physical activity [25]. Alcohol consumption was assessed by asking individuals how many days in the past month they had consumed alcohol. This number was then multiplied by their self-reported daily alcohol intake to calculate the weekly consumption, expressed in grams alcohol per week [23].

Information on current medications was collected through an interview with a trained nurse and included detailed information on the name of the medication, indication, dosage (expressed as dose per medication), administration period, and dose per administration occasion. Antihypertensive medication was defined as the prescription of at least one of the following drug classes: Angiotensin-Converting Enzyme Inhibitors (ACEi), Angiotensin II Receptor Blockers (ARB), Alpha-Blockers, Beta-Blockers (BB), Calcium Channel Blockers (CCB), Thiazide Diuretics, Aldosterone Antagonists or Loop Diuretics.

#### Statistics

The primary exposure variables were the arterial elasticity as C1 and C2. Each exposure was analysed as a standardized z-score (per 1 SD increase) and as a categorical variable using sex-specific quartiles. The primary outcome was left ventricular ejection fraction (EF), analysed as a continuous variable. Associations between C1 and C2 with EF (separately) were assessed using multivariable linear regression in separate models. To compare adjusted mean EF across levels of C2 quartiles (as categorical predictors), instead of continuous C2, were used in linear regression models. Left ventricular (LV) mass was analysed both as a continuous outcome (linear regression) and as a binary outcome indicating left ventricular hypertrophy (LVH) based on established sex-specific thresholds, using logistic regression. Analyses using sex-specific quartiles were considered secondary and were performed to aid clinical interpretability; the primary inference was based on the continuous z-score models.

##### Covariates and statistical models

Potential confounders were defined a priori as variables expected to be associated with both arterial elasticity indices and cardiac outcomes. Associations between C1, C2 and echocardiographic measures were evaluated using four sequential multivariable models: *Model 1*: adjusted for sex, age, and heart rate. *Model 2*: additionally adjusted for diabetes mellitus, LDL-cholesterol, BMI, and antihypertensive medication use (yes/no). (to adjust for pharmacologic associations with elasticity and cardiac function [7, 26]). *Model 3* further adjusted for lifestyle factors: current smoking, alcohol intake (non-user vs consumer), and leisure-time physical activity (LTPA). *Model 4* additionally adjusted for SBP.

We tested potential effect modification by sex, hypertension status, and history of major cardiovascular disease by including interaction terms between these variables and C1 or C2. Additional sensitivity analyses were performed by replacing SBP with pulse pressure. In addition, we repeated the analyses using the number of prescribed antihypertensive medications instead of antihypertensive medication use as a covariate.

#### Use of Artificial Intelligence

AI tools were used for language support, editing, and translation, where applicable. The authors remain fully responsible for the content of the manuscript’s scientific accuracy.

## Results

### Study characteristics

The final study population consisted of 991 participants (488, 49% men) with complete data on C2 and cardiac function. The characteristics of this population are shown in *Table 1*. Men had higher systolic and diastolic blood pressure and fasting plasma glucose, HOMA-IR, and LDL cholesterol. Smoking prevalence was about 19%, and more women were non-drinkers (28% vs. 14%,). Low levels of self-reported leisure time physical activity was common (66%), especially among women (72% vs. 62%). In line with observations in previous studies, C1 and C2 were higher in men than in women. LVMI was higher in men, while ejection fraction and LVH prevalence were similar across sexes. Ejection fraction and LVH was similar across sexes *(Table 1)*.

**Table 1.** Characteristics of the study population with data on arterial elasticity indices (C1 and C2) and cardiac function.

|  | **All (n=991)** | **Men (n=488)** | **Women (n=503)** |
| --- | --- | --- | --- |
| Age, years<sup>a</sup> | 50.6 (11.9) | 50.8 (12.2) | 50.5 (11.6) |
| SBP, mmHg<sup>a</sup> | 125 (18) | 127 (16) | 123 (18) |
| DBP, mmHg<sup>a</sup> | 71 (10) | 73 (10) | 70 (10) |
| Pulse, bpm<sup>a</sup> | 63 (8) | 63 (9) | 64 (8) |
| BMI, kg/m<sup>2a</sup> | 27.0 (4.3) | 26.9 (3.4) | 27.1 (5.1) |
| WHR<sup>a</sup> | 0.9 (0.1) | 1.0 (0.1) | 0.8 (0.1) |
| BSA, m<sup>2a</sup> | 1.9 (0.2) | 2.0 (0.1) | 1.8 (0.2) |
| Fasting glucose, mmol/L<sup>a</sup> | 5.5 (1.3) | 5.6 (1.1) | 5.4 (1.6) |
| HOMA-IR, median (IQR) | 1.2 (0.7–1.9) | 1.3 (0.8–2.0) | 1.1 (0.7–1.7) |
| LDL cholesterol, mmol/L<sup>a</sup> | 3.3 (0.9) | 3.4 (0.9) | 3.3 (0.9) |
| Total hypertension, n (%) | 184 (19) | 91 (19) | 93 (19) |
| Total DM, n (%) | 69 (7) | 34 (7) | 35 (7) |
| Prescribed antihyperlipidemic medication, n (%) | 40 (4) | 21 (4) | 19 (4) |
| Prescribed antihypertensive medication, n (%) | 128 (13) | 58 (12) | 70 (14) |
| Current smoker, n (%) | 184 (19) | 81 (17) | 103 (21) |
| Non-drinker, n (%) | 209 (22) | 68 (14) | 141 (28) |
| Low-level of physical activity (LTPA: 1 and 2), n (%) | 657 (66) | 298 (62) | 359 (72) |
| C2, mL/mmHg×100<sup>a</sup> | 6.9 (3.5) | 7.6 (3.5) | 6.2 (3.3) |
| C1, mL/mmHg×10<sup>a</sup> | 16.0 (5.1) | 17.4 (5.2) | 14.8 (4.7) |
| Ejection fraction, %<sup>a</sup> | 73.4 (8.5) | 73.4 (8.5) | 74.2 (8.6) |
| LVMI, g/m<sup>2.7</sup> <sup>a</sup> | 37.4 (10.7) | 38.5 (10.6) | 36.4 (10.6) |
| LVH, n (%) | 160 (16.1) | 80 (16.4) | 80 (15.9) |

<sup>a</sup>: data are presented as mean (SD). SBP: systolic blood pressure; DBP: diastolic blood pressure; BMI: body mass index; BSA: body surface area; HOMA-IR: homeostatic model assessment for insulin resistance; LDL: low-density lipoprotein; DM: diabetes mellitus; C2: small artery elasticity index; C1: large artery elasticity index; LTPA: leisure-time physical activity; LVMI: left ventricular mass index; LVH: left ventricular hypertrophy.

### Associations of arterial elasticity with ejection fraction

Higher C2 was associated with higher ejection fraction. In the SBP-adjusted model, a 1-SD higher C2 was associated with 1.2 percentage points higher EF (β = 1.2, 95% CI: 0.5–1.9; p < 0.001) (*Table 2*). Similar results were observed when antihypertensive medication use was replaced by the number of prescribed antihypertensive medications (β = 1.2, 95% CI: 0.5–1.9; p < 0.001), and when systolic blood pressure was replaced by pulse pressure (β = 1.4, 95% CI: 0.7–2.1; p < 0.001). In the SBP-adjusted model, EF was highest in Q4 and lowest in Q1 (mean ± SE: 75.3 ± 0.6% vs. 72.6 ± 0.6%). EF was significantly higher in Q4 than in Q1 (mean difference: 2.7 percentage points, 95% CI: 0.8 to 4.6; p = 0.005) (*Figure 1.*). Similar associations were observed for the association between C1 and EF (Model 3: β = 0.6, 95% CI: 0.03– 1.2; p = 0.039). However, this association was no longer statistically significant after adjustment for SBP in the final model (β = 0.3, 95% CI: −0.4 to 0.9; p = 0.416). In the sensitivity analysis replacing SBP with pulse pressure, the estimate was larger but still not statistically significant (β = 0.6, 95% CI: −0.01 to 1.2; p = 0.055).

**Figure 1.**
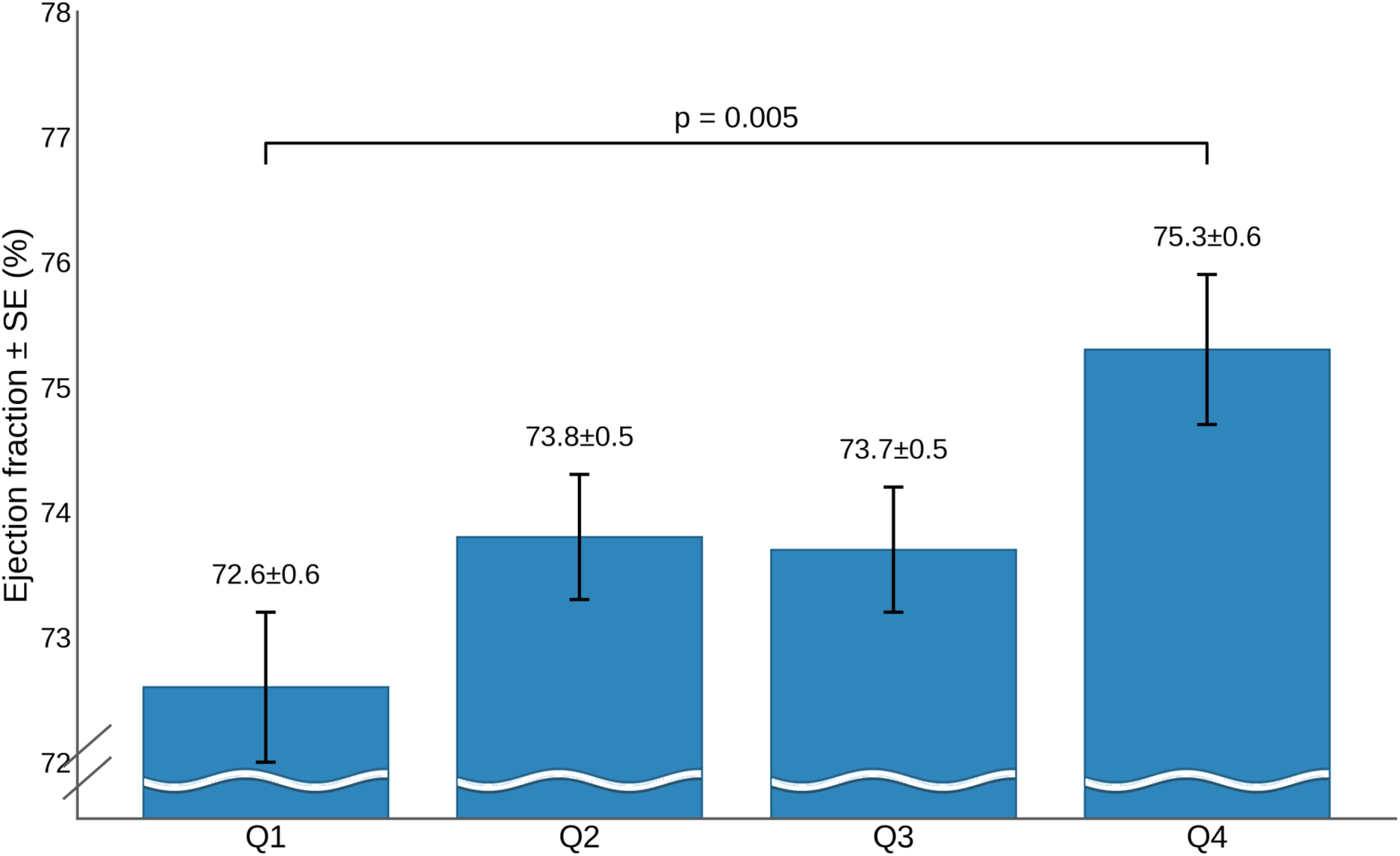
Associations of sex-specific quartiles of Small artery elasticity index (C2) with Ejections fraction (EF). N=991. Adjusted for age, sex, heart rate, current diagnosis of diabetes mellitus, LDL (Low-Density Lipoprotein) - cholesterol, body mass index, use of antihypertensive medicines, current smoking, alcohol intake, leisure time physical activity and systolic blood pressure

**Table 2.** Associations Between Small Artery Elasticity Index (C2) and Systolic Functions in the Vara-Skövde Cohort (N=991)

|  | Ejection fraction (EF) |  |  |
| --- | --- | --- | --- |
| | $\beta$ | 95% CI | p-value |
| Model 1 | 1.6 | 1.0-2.3 | <0.001 |
| Model 2 | 1.4 | 0.8-2.1 | <0.001 |
| Model 3 | 1.4 | 0.8-2.1 | <0.001 |
| Model 4 | 1.2 | 0.5-1.9 | <0.001 |

The results are based linear regressions with key exposure variable as small artery elasticity index (C2), and outcome as ejection fraction (EF). The results are expressed as per 1 standard deviation increase in C2. Model 1 was adjusted for age, heart rate, and sex; Model 2 additionally for diabetes mellitus, LDL cholesterol, body mass index (BMI), and antihypertensive medication use (yes/no).; Model 3 further for current smoking, alcohol intake, and leisure-time physical activity (LTPA); and Model 4 additionally for systolic blood pressure (SBP). $\beta$ denotes the unstandardized regression coefficient; CI, confidence interval.

### Associations of arterial elasticity with left ventricular mass index and left ventricular hypertrophy

Higher small artery elasticity index (C2) was associated with lower left ventricular mass index (LVMI). In Model 3, each standard deviation higher C2 was associated with 1.2 units lower LVMI (β: −1.2, 95% CI: −2.0- −0.5, p = 0.001). However, the association was no longer significant in the final model (β: −0.3, 95% CI: −1.0- 0.5, p = 0.489). Similarly, higher C1 was associated with lower LVMI in Model 3 in (β = −1.7, 95% CI: −2.4 to −1.0; p < 0.001) but the association was no longer significant in Model 4 (β = −0.6, 95% CI: −1.3 - 0.1; p = 0.116).

Higher C2 was associated with lower odds of LVH in Model 3, although this association was attenuated and no longer statistically significant after additional adjustment for SBP (Model 3: OR 0.58, 95% CI 0.41–0.8; p=0.001; Model 4: OR 0.81, 95% CI 0.58–1.15; p = 0.245). In the sensitivity analysis using pulse pressure instead of SBP, the association remained significant (OR 0.71, 95% CI 0.51–0.99; p = 0.047).

In contrast, a higher C1 was consistently associated with lower odds of LVH, including after adjustment for SBP (Model 3: OR 0.47, 95% CI 0.35–0.63; p < 0.001; Model 4: OR 0.63, 95% CI 0.46–0.87; p = 0.005) *(Table 3).* The findings remained consistent in sensitivity analyses replacing SBP with pulse pressure in the fully adjusted model (OR 0.59, 95% CI 0.44–0.81; p < 0.001) and antihypertensive medication use with the number of prescribed antihypertensive medications (OR 0.61, 95% CI 0.44–0.84; p = 0.003).

**Table 3.** Associations Between Arterial Elasticity Indices (C1, C2) and Left Ventricular Hypertrophy (LVH) in the Vara–Skövde Cohort (N=991)

|  | Large Artery Elasticity Index (C1) |  |  | Small Artery Elasticity Index (C2) |  |  |
| --- | --- | --- | --- | --- | --- | --- |
|  | OR | 95% CI | p-value | OR | 95% CI | p-value |
| Model 1 | 0.57 | 0.42- 0.75 | <0.001 | 0.67 | 0.51- 0.89 | 0.005 |
| Model 2 | 0.47 | 0.35- 0.63 | <0.001 | 0.58 | 0.42- 0.79 | <0.001 |
| Model 3 | 0.47 | 0.35- 0.63 | <0.001 | 0.58 | 0.41- 0.8 | 0.001 |
| Model 4 | 0.63 | 0.46- 0.87 | 0.005 | 0.81 | 0.58- 1.15 | 0.245 |

The results are based logistic regression for associations of C1 and C2 with LVH. The results are expressed per 1 standard deviation increase in each index. Model 1 was adjusted for age, heart rate, and sex; Model 2 additionally is adjusted for diabetes mellitus, LDL cholesterol, body mass index (BMI), and use of prescribed antihypertensive medications; Model 3 further is adjusted for current smoking, alcohol intake, and leisure-time physical activity (LTPA); and Model 4 additionally for systolic blood pressure (SBP). OR: odds ratio; CI: confidence interval.

Sensitivity analyses were performed to assess the robustness of the findings. Although an interaction between C2 and sex was observed for EF (p = 0.04), it was no longer significant after also including interactions between sex and age and heart rate (p = 0.965). There was also no evidence of interactions between arterial elasticity and sex when the outcome was left ventricular mass (p=0.3). Thus, sex-stratified analyses were not performed.

## Discussion

### Main findings

Our study demonstrates a consistent association between arterial elasticity and left-ventricular systolic performance and morphology. Higher Small Artery Elasticity Index (C2) was significantly associated with higher ejection fraction (EF) after multivariable adjustment. Moreover, Large Artery Elasticity Index (C1) was inversely associated with left-ventricular hypertrophy (LVH) even after adjusting for systolic blood pressure, suggesting that C1 captures afterload components not completely reflected by brachial SBP, most plausibly central reservoir compliance and related pulsatile load [27, 28].

To our knowledge, these associations have not previously been reported using diastolic pulse wave methodology. Prior work has consistently linked arterial stiffening and pulsatile hemodynamics to adverse LV remodelling and heart-failure risk. In a hypertensive cohort, increase in carotid-femoral pulse wave velocity (cfPWV) and central pulse pressure were associated with increase in LV mass and concentric remodelling, [29]. In the Multi-Ethnic Study of Atherosclerosis (MESA), greater arterial wave reflection magnitude predicted heart failure [13]. Also in the MESA cohort, Windkessel-derived indices summarizing the diastolic pressure decay (PTC1/PTC2) were also associated with incident heart failure [12]. Consistent with these observations, a recent systematic review and meta-analysis reported significantly higher PWV including cfPWV and brachial–ankle pulse wave velocity (baPWV), in patients with HT compared with controls [10].

Against this background, our findings extend the literature by showing that elasticity indices measured by diastolic pulse wave capture complementary ventricular–vascular phenotypes: C2 shows a significant association with systolic performance beyond brachial SBP, whereas C1 shows a clear inverse association with left ventricular hypertrophy even after SBP adjustment. The persistence of the associations after replacing systolic blood pressure with pulse pressure suggests that the findings were robust to the choice of blood-pressure adjustment variable.

### Small-artery elasticity and left-ventricular systolic cardiac function

C2 reflects resistance-vessel tone and systemic microvascular function. Higher small-artery elasticity may reduce wave reflections and thereby reduce left ventricular afterload. C2 may also capture neurohormonal and endothelial influences on vascular tone: increased sympathetic and renin–angiotensin–aldosterone system activity promotes peripheral vasoconstriction and would be expected to lower C2, while endothelial dysfunction and microvascular impairment are similarly associated with reduced C2. In phenotypes with reduced systolic function, higher arterial stiffness can worsen haemodynamics by raising afterload and reducing arterial buffering [30].

### Artery elasticity and left-ventricular mass

Both C2 and C1 were inversely associated with LVMI, but these associations were attenuated after adjustment for SBP, suggesting that they may be mediated by blood pressure a finding that is consistent with previous reports [27]. This is similar to earlier studies showing inverse relationships between C2 respective C1 and LVMI, although those analyses were limited by less extensive adjustment and were conducted in selected hypertensive population [31]. Similarly, the significant inverse association between C2 and LVH was no longer evident after SBP adjustment, whereas C1 remained inversely associated with LVH. This supports the concept that C1 reflects central conduit-artery compliance and more chronic structural vascular alterations, which may be more closely related to ventricular remodelling, particularly at earlier stages of disease [28, 30]. The loss of the C2–LVH association after SBP adjustment further suggests that remodelling is driven mainly by large-artery stiffness and cumulative blood pressure load, with possible additional contributions from cardiometabolic factors such as insulin resistance and diabetes mellitus [32]. In a high cardiovascular-risk cohort, measures more closely reflecting central haemodynamics, including cfPWV and central pulse pressure, were more strongly associated with incident heart failure than peripheral PWV assessed by baPWV [33]. Because the present study was cross-sectional, these findings should be interpreted as consistent with associations with cardiac remodelling rather than evidence of incident heart failure risk.

### Strengths and limitations

Several limitations should be considered. First, the cross-sectional design limits causal inference, and the observed associations should not be interpreted as evidence of causal relationships. Second, systolic blood pressure is part of the algorithm for calculating C1 and C2. Therefore, adjusting for levels of SBP in regression models can complicate interpretation. Blood pressure is a well-known risk factor for deterioration of heart function and morphology. Thus, our aim was to investigate whether these associations remained significant beyond blood pressure adjustments. Therefore, we chose to present Model 4 as a specific model. However, brachial SBP and pulse pressure are imperfect surrogates of central haemodynamic load. The persistence of the C1–LVH association after adjustment for either SBP or pulse pressure suggests that C1 may capture vascular properties not fully reflected by conventional brachial blood pressure indices.

Third, C1 and C2 are derived from a Windkessel-based pulse contour algorithm that incorporates device-estimated haemodynamic variables, including estimated cardiac output through systemic vascular resistance [34]. For this reason, the association between C1/C2 and cardiac output was not analysed in the present study. This would have added important information exploring the association between vascular elasticity and cardiac systolic function.

Several conditions other than hypertension may contribute to increased left ventricular mass. We therefore adjusted for systolic blood pressure (SBP) and a diagnosis of diabetes mellitus (DM). Diabetes was included as an important confounder because of its established associations with myocardial stiffness, left ventricular hypertrophy, and fibrosis [32]. We used clinically defined diabetes rather than HOMA-IR, as it is a more stable and clinically relevant measure. SBP was retained both to align with previous literature [12, 33] and to assess whether C1 and C2 provide information beyond SBP and traditional risk factors. However, we lacked data on genetic variants associated with cardiomyopathy and had no information on aortic stenosis or cardiac amyloidosis, both of which may have influenced left ventricular mass and introduced residual confounding.

The main strength of this study is the observation in a large community-based cohort without clinical heart failure, which permitted investigation of associations between vascular function and ventricular structure and function at an early stage, before the development of clinical signs of disease. Moreover, the meticulous characterization of this cohort allowed adjustment for several relevant confounders, thereby adding new knowledge to the existing literature.

## Conclusions

Hypertension is the leading modifiable contributor to heart failure. In this study, higher Small Artery Elasticity Index (C2) was associated with greater systolic performance, as reflected by higher ejection fraction beyond blood pressure, while the Large Artery Elasticity Index (C1) was inversely associated with LVH risk. These findings suggest that early vascular assessment with diastolic pulse wave analysis may provide a non-invasive approach to identify subclinical cardiovascular alterations before overt disease develops. The magnitude of observed associations with EF were modest, and the measures reflect subtle physiological variation rather than clinically overt systolic dysfunction. Their main relevance may therefore be pathophysiological and preventive, by improving understanding of early ventriculo-arterial interactions and subclinical cardiovascular risk, rather than immediate individual-level clinical decision-making.

## Data Availability

The datasets analyzed during the current study are not publicly available due to ethical and legal restrictions related to participant confidentiality. Data access is restricted by the terms of the ethical approval granted by the Ethics Committee at the University of Gothenburg, Sweden, and data sharing is not permitted under current regulations. Researchers may request access to anonymized data by contacting the corresponding author, subject to approval by the ethics committee and institutional data protection policies.

## Acknowledgements

We want to thank all participants and study nurses for their participation and devotion in this study.

## Nonstandard Abbreviations and Acronyms

ACEi: angiotensin-converting enzyme inhibitor
ARB: angiotensin II receptor blocker
baPWV: brachial-ankle pulse wave velocity
BB: beta-blocker
C1 or LAEI: large artery elasticity index
C2 or SAEI: small artery elasticity index
cfPWV: carotid-femoral pulse wave velocity
CCB: calcium channel blocker
GLM: generalized linear model
HFmrEF: heart failure with mildly reduced ejection fraction
HFpEF: heart failure with preserved ejection fraction
HFrEF: heart failure with reduced ejection fraction
JNC-7: Seventh Report of the Joint National Committee on Prevention, Detection, Evaluation, and Treatment of High Blood Pressure
LTPA: leisure-time physical activity
LVH: left ventricular hypertrophy
LVMI: left ventricular mass index
OGTT: oral glucose tolerance test
MESA: Multi-Ethnic Study of Atherosclerosis
SBP: systolic blood pressure

## References

1. Ran, J., et al., Global, regional, and national burden of heart failure and its underlying causes, 1990–2021: results from the global burden of disease study 2021. Biomarker Research, 2025. 13(1): p. 16.

2. Harada, T., et al., Echocardiography in the diagnostic evaluation and phenotyping of heart failure with preserved ejection fraction. Journal of Cardiology, 2022. 79(6): p. 679–690.

3. Youn, J.C., Y. Ahn, and H.O. Jung, Pathophysiology of Heart Failure with Preserved Ejection Fraction. Heart Fail Clin, 2021. 17(3): p. 327–335.

4. Herzog, M.J., et al., Arterial stiffness and vascular aging: mechanisms, prevention, and therapy. Signal Transduction and Targeted Therapy, 2025. 10(1): p. 282.

5. Vlachopoulos, C., K. Aznaouridis, and C. Stefanadis, Prediction of cardiovascular events and all-cause mortality with arterial stiffness: a systematic review and meta-analysis. J Am Coll Cardiol, 2010. 55(13): p. 1318–27.

6. Ben-Shlomo, Y., et al., Aortic pulse wave velocity improves cardiovascular event prediction: an individual participant meta-analysis of prospective observational data from 17,635 subjects. J Am Coll Cardiol, 2014. 63(7): p. 636–646.

7. Chirinos, J.A., et al., Large-Artery Stiffness in Health and Disease: JACC State-of-the-Art Review. J Am Coll Cardiol, 2019. 74(9): p. 1237–1263.

8. Szalo, G., et al., Impaired artery elasticity predicts cardiovascular morbidity and mortality- A longitudinal study in the Vara-Skovde Cohort. J Hum Hypertens, 2023.

9. Saz-Lara, A., et al., Association Between Arterial Stiffness and Blood Pressure Progression With Incident Hypertension: A Systematic Review and Meta-Analysis. Frontiers in Cardiovascular Medicine, 2022. Volume 9 - 2022.

10. Esmaeili, Z., et al., The association between pulse wave velocity and heart failure: a systematic review and meta-analysis. Front Cardiovasc Med, 2024. 11: p. 1435677.

11. Duprez, D.A., Arterial stiffness/elasticity in the contribution to progression of heart failure. Heart Fail Clin, 2012. 8(1): p. 135–41.

12. Brumback, L.C., et al., The association between indices of blood pressure waveforms (PTC1 and PTC2) and incident heart failure. J Hypertens, 2021. 39(4): p. 661–666.

13. Chirinos, J.A., et al., Arterial wave reflections and incident cardiovascular events and heart failure: MESA (Multiethnic Study of Atherosclerosis). J Am Coll Cardiol, 2012. 60(21): p. 2170–7.

14. Chobanian, A.V., et al., Seventh report of the Joint National Committee on Prevention, Detection, Evaluation, and Treatment of High Blood Pressure. Hypertension, 2003. 42(6): p. 1206–52.

15. Alberti, K.G. and P.Z. Zimmet, Definition, diagnosis and classification of diabetes mellitus and its complications. Part 1: diagnosis and classification of diabetes mellitus provisional report of a WHO consultation. Diabet Med, 1998. 15(7): p. 539–53.

16. Daka, B., et al., Association between self-reported alcohol consumption and diastolic dysfunction: a cross-sectional study. BMJ Open, 2023. 13(10): p. e069937.

17. Ingelsson, E., et al., The PPARGC1A Gly482Ser polymorphism is associated with left ventricular diastolic dysfunction in men. BMC Cardiovasc Disord, 2008. 8: p. 37.

18. Lang, R.M., et al., Recommendations for chamber quantification: a report from the American Society of Echocardiography’s Guidelines and Standards Committee and the Chamber Quantification Writing Group, developed in conjunction with the European Association of Echocardiography, a branch of the European Society of Cardiology. J Am Soc Echocardiogr, 2005. 18(12): p. 1440–63.

19. Jarnert, C., et al., Doppler tissue imaging in congestive heart failure patients due to diastolic or systolic dysfunction: a comparison with Doppler echocardiography and the atrio-ventricular plane displacement technique. Eur J Heart Fail, 2000. 2(2): p. 151–60.

20. Gudmundsson, P., et al., Visually estimated left ventricular ejection fraction by echocardiography is closely correlated with formal quantitative methods. Int J Cardiol, 2005. 101(2): p. 209–12.

21. Krauser, D.G. and R.B. Devereux, Ventricular hypertrophy and hypertension: prognostic elements and implications for management. Herz, 2006. 31(4): p. 305–16.

22. de Simone, G., et al., Metabolic syndrome and left ventricular hypertrophy in the prediction of cardiovascular events: the Strong Heart Study. Nutr Metab Cardiovasc Dis, 2009. 19(2): p. 98–104.

23. Goransson, M. and B.S. Hanson, How much can data on days with heavy drinking decrease the underestimation of true alcohol consumption? J Stud Alcohol, 1994. 55(6): p. 695–700.

24. Grimby, G., et al., The “Saltin-Grimby Physical Activity Level Scale” and its application to health research. Scand J Med Sci Sports, 2015. 25 Suppl 4: p. 119–25.

25. Lochen ML, R.K., The Tromso study: Physical fitness, self-reported physical activity, and their relationship to other coronary risk factors. J Epidemiol Community Health 1992. 46: p. 103–7.

26. Cavero-Redondo, I., et al., Comparative effect of antihypertensive drugs in improving arterial stiffness in adults with hypertension (RIGIPREV study). A network meta-analysis. Front Pharmacol, 2023. 14: p. 1225795.

27. Agbaje, A.O., J.P. Zachariah, and T.P. Tuomainen, Arterial stiffness but not carotid intima-media thickness progression precedes premature structural and functional cardiac damage in youth: A 7-year temporal and mediation longitudinal study. Atherosclerosis, 2023. 380: p. 117197.

28. Zamani, P., et al., Resistive and pulsatile arterial load as predictors of left ventricular mass and geometry: the multi-ethnic study of atherosclerosis. Hypertension, 2015. 65(1): p. 85–92.

29. Tan, J., et al., Aortic Pulse Wave Velocity is Associated with Measures of Subclinical Target Organ Damage in Patients with Mild Hypertension. Cell biochemistry and biophysics, 2014. 70(1): p. 167–171.

30. Chirinos, J.A. and N. Sweitzer, Ventricular–Arterial Coupling in Chronic Heart Failure. Cardiac Failure Review 2017;3(1):12–8., 2017.

31. Cernes, R., et al., Relation of arterial properties to left ventricular hypertrophy in hypertensive adults: focus on gender-related differences. Angiology, 2010. 61(5): p. 510–515.

32. Jia, G., M.A. Hill, and J.R. Sowers, Diabetic Cardiomyopathy. Circulation Research, 2018. 122(4): p. 624–638.

33. Lee, C.J., et al., Comparison of the Association Between Arterial Stiffness Indices and Heart Failure in Patients With High Cardiovascular Risk: A Retrospective Study. Frontiers in Cardiovascular Medicine, 2021. Volume 8 - 2021.

34. Cohn, J.N., et al., Noninvasive pulse wave analysis for the early detection of vascular disease. Hypertension, 1995. 26(3): p. 503–8.

